# Use of rapid molecular tuberculosis diagnostics across Brazil’s incarcerated population

**DOI:** 10.1101/2021.11.30.21266839

**Authors:** Elinor B. Fajer, Fernanda Dockhorn Costa, Daniele Maria Pelissari, Fredi Alexander Diaz Quijano, Artemir Coelho de Brito, Eunice Atsuko Totumi Cunha, Julio Croda, Jason R. Andrews, Katharine S. Walter

## Abstract

**Background:** Incarcerated individuals in Brazil are at high-risk of tuberculosis (TB), but their access to World Health Organization recommended diagnostics is poorly understood.

**Methods:** We conducted a retrospective cross-sectional study of newly diagnosed TB cases in Brazil’s notifiable disease registry, which includes information on diagnostic tests performed, from January 2015 through December 2018. We quantified reported use of TB diagnostics across incarcerated and non-incarcerated populations and tested for municipality-level factors associated with diagnostic usage among the incarcerated population with generalized linear regression.

**Results:** Between 2015 and 2018, 258,014 individuals were newly diagnosed with TB, including 27,400 (10.6%) incarcerated individuals. Among these, 27.5% had an Xpert MTB/RIF test reported; 71.5% had sputum smear; 34.1% had culture; 70.9% had chest radiography. Xpert MTB/RIF use was greater among incarcerated than non-incarcerated individuals (36.2% vs 26.5%, p<0.001). However, we found spatial heterogeneity in state-level use of both Xpert MTB/RIF (range: 4.7-72.4% cases diagnosed) and chest radiography (range: 11.7-88.4%) in prisons. We identified seven municipalities with large incarcerated populations (>5000) with rates of Xpert MTB/RIF usage below the national average in incarcerated individuals.

**Conclusion:** Prioritizing expansion of rapid molecular diagnostics in prisons, particularly in regions with limited current usage of molecular diagnostics, will be an essential component of TB control.

## INTRODUCTION

Tuberculosis (TB) is one of the leading causes of death by infectious disease worldwide^1^. However, there remains a large population of undiagnosed TB cases, which the COVID-19 pandemic has further expanded^1^. In 2020, of the estimated 9.9 million globally incident TB cases, only 5.8 million cases were reported^1^. Reducing this diagnostic gap through early diagnosis of disease and rapid detection of drug resistance is critical for both managing individual outcomes and for preventing onwards transmission of TB^1–3^.

As part of the End TB Strategy, the World Health Organization (WHO) recommends that all individuals with symptoms of TB should have access to WHO-endorsed molecular rapid diagnostic tests^4^. Rapid molecular diagnostic tests such as Xpert MTB/RIF have high sensitivity (89%) and specificity (99%) in detecting *M. tuberculosis* when compared to a microbiological reference standard and a low limit of detection^5,6^, enabling detection of *M. tuberculosis* in paucibacillary samples. Further, automated molecular diagnostics like Xpert MTB/RIF do not require biosafety level three facilities, extensive laboratory capacity, or long follow-up periods, as do many other diagnostics. However, in many high-incidence countries, sputum smear microscopy remains the most used diagnostic test, and culture remains the gold standard for TB positivity.

Expanding access to rapid molecular TB diagnostics is an urgent priority for vulnerable populations such as incarcerated individuals, who globally have a TB incidence ten times that of the general population^7^. In the Americas, TB notifications are increasingly concentrated within prisons^8,9^, including in Brazil, where 12.2% of the country’s notified TB cases occurred among incarcerated individuals in 2019^11^. The conditions of detention put individuals at high risk of TB infection and often reduce their contact with healthcare systems, resulting in a lower probability of early diagnosis and treatment^9,12^. Higher TB levels across incarcerated populations not only affect these groups of people directly, but also have impacts at the population level due to amplifying factors such as spillover^13–15^.

Early diagnosis and treatment is critical for reducing the burden of TB in prisons, and the speed, accessibility, and cost effectiveness of Xpert MTB/RIF makes it especially suitable in prison settings^16^. Modeling studies have found that active screenings with Xpert MTB/RIF can significantly reduce the burden of TB among incarcerated populations^13,17–19^. While the Brazil National Tuberculosis Program recently endorsed the use of Xpert MTB/RIF, the usage of rapid molecular diagnostics in prisons has not yet been explored. Additionally, as studies have supported chest radiography use for mass screenings in prison settings^17^, the usage of this diagnostic across Brazil’s incarcerated population is also of interest.

Here, we analyzed Brazil’s national notifiable disease registry to quantify the use of Xpert MTB/RIF and chest radiography across Brazil’s incarcerated population and to identify critical gaps in the usage of recommended diagnostics.

## METHODS

### Study design and data sources

We conducted a retrospective cohort study of newly diagnosed tuberculosis cases reported to Brazil’s national notifiable disease system, Sistema de Informação de Agravos de Notificação (SINAN), from January 2015 through December 2018. In Brazil, individual tuberculosis diagnoses are compulsorily reported in SINAN^20^. SINAN includes patient clinical and sociodemographic information, incarceration status at the time of TB notification, TB diagnostic test results, and TB treatment outcomes. We restricted the study period to between 2015 and 2018 as Xpert MTB/RIF implementation began in late 2014, and data for years following 2018 were incomplete at the time of the study. We excluded cases with unknown incarceration status or of patients below 18 years of age, as they make up a small proportion of the incarcerated population in Brazil.

Patients may receive more than one diagnostic test to confirm their TB diagnosis. SINAN reports diagnostic results by testing modality, including Xpert MTB/RIF, chest radiography, sputum smear, and culture. Xpert MTB/RIF test outcomes are reported as *M. tuberculosis* detected, sensitive to Rifampicin; *M. tuberculosis* detected, resistant to Rifampicin; *M. tuberculosis* not detected; inconclusive test; or not performed. Xpert MTB/RIF G4 was primarily used during our study period, though Brazil introduced Xpert MTB/RIF Ultra across the country’s laboratory network in 2019. Chest radiography test outcomes are reported as suspect, normal, other pathology, or not performed; sputum smear outcomes are reported as positive, negative, not performed, or not applicable; and culture outcomes are reported as positive, negative, in-progress, or not performed. For each individual diagnostic test, we classified individuals with positive, negative, or in-progress reported results as having received that test (test usage), and individuals with tests marked not performed or missing information as having not received that diagnostic test (no test usage). At the municipality level, we defined test use as the proportion of cases diagnosed with each respective diagnostic tool.

In addition to the individual level variables extracted from SINAN, we collected municipality-level covariates, including average per capita household income and GINI coefficient, from the Human Development Atlas^21^. The Human Development Atlas collects data each decade; therefore, we used data from the most recent report (2010). We obtained incarcerated population size by municipality from Brazil’s National Prison Information Survey^22^ and data for total municipality population density from the Brazilian Institute of Geography and Statistics^23^.

### Statistical Analysis

Data analysis was conducted using R (version 3.6.3) and R Studio (version 1.3.1093). We first compared the proportions of patients diagnosed with Xpert MTB/RIF, chest radiography, sputum smear, and culture between incarcerated and non-incarcerated populations with two-proportion z-tests. A multiple samples proportions test was performed to assess the increasing trend in Xpert MTB/RIF use over the study period.

We calculated use of the four testing modalities at state and municipality levels based on individual-level data from SINAN and the location of patients’ diagnoses. Chi-square tests for equality of proportions were run to assess the significance of state-level diagnostic use heterogeneity. An ‘empty’ multilevel regression^24^ was run with state or municipality random effects respectively and no other predictor variables. These models were used to estimate the intraclass correlation coefficients (ICC) for municipality and state-level clustering, a measure of the proportion of total variability as determined by cluster membership.

We used mixed effects binomial regression to identify covariates associated with use of Xpert MTB/RIF and chest radiography among the incarcerated population. We included municipality population density, GINI coefficient, incarcerated population size, average per capita household income, and year (2015 through 2018) as fixed effects, and municipality of TB diagnosis as a random effect. The population density, incarcerated population size, and average per capita household income variables were on a log scale. The GINI coefficient was rescaled by a factor of ten for to optimize model fitting.

We investigated gaps in diagnostic use for all Brazil municipalities with incarcerated populations of at least 5000 individuals and with at least 5 TB cases from 2015 through 2018.

### Ethics Statement

This study was approved by the Institutional Review Board at Stanford University (Protocol #50466).

## RESULTS

In Brazil between 2015 and 2018, there were 284,415 newly diagnosed TB cases reported to Brazil’s Notifiable Diseases Information System (SINAN), of which 258,014 were among patients over 18 years of age and with information on incarceration status. Of these notified cases, 10.6% (27,400 of 258,014 cases) occurred among incarcerated individuals and 89.4% (230,614 of 258,014) were among the general population. We found overall Xpert MTB/RIF use of 27.5% (70,988 of 258,014), chest radiography use of 70.9% (183,058 of 258,014), sputum smear use of 71.5% (184,509 of 258,014), and culture use of 34.1% (87,905 of 258,014), with significant overlap in populations receiving different diagnostic tests (Fig. 1). TB patients received a mean of 2.04 diagnostic tests, with the greatest overlap of 27.4% of patients receiving both chest radiography and sputum smear.

**Figure 1:**
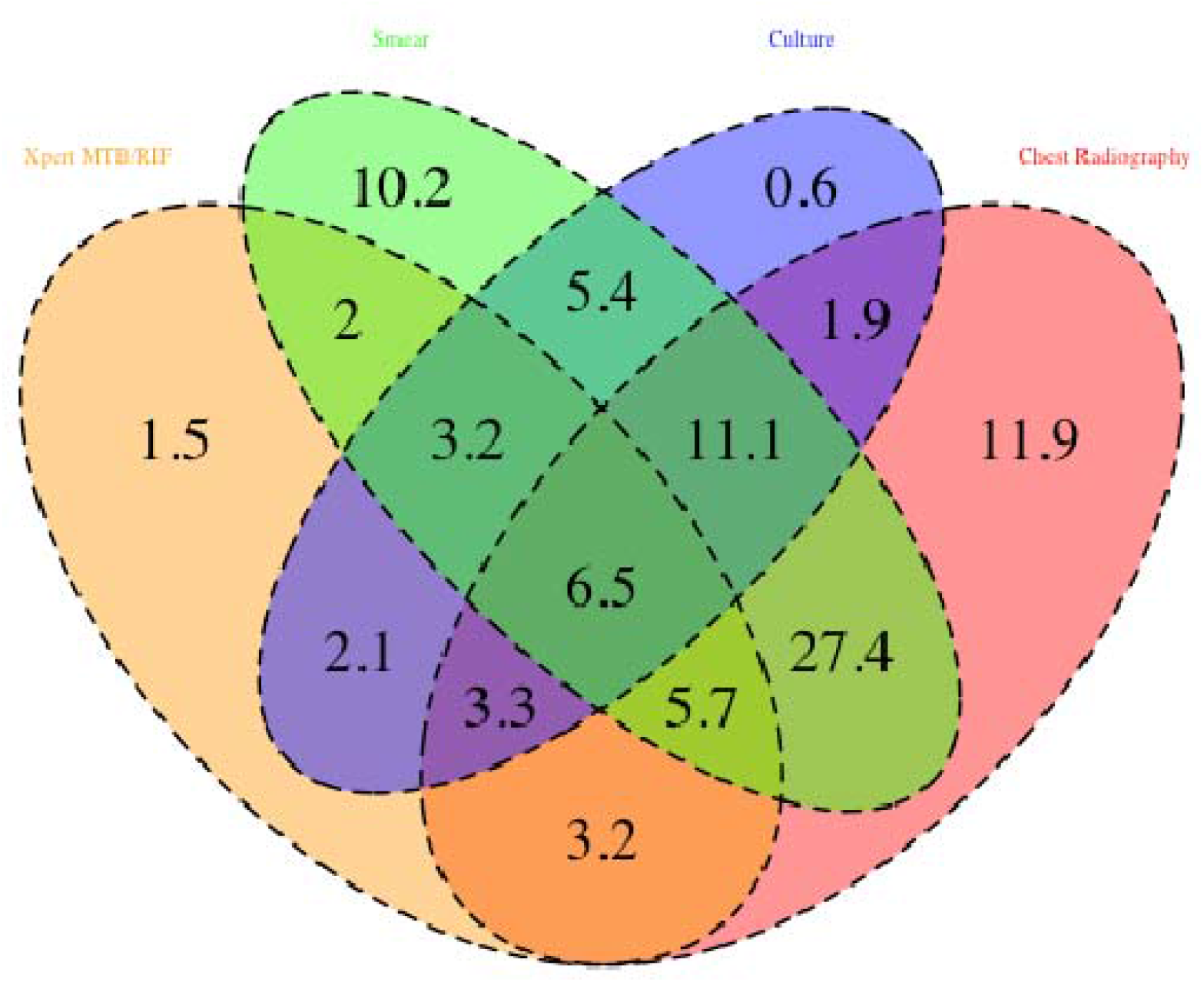
Venn diagram of TB diagnostic use. Percentage of newly diagnosed TB cases with diagnostic test results reported for 1) Xpert MTB/RIF, 2) Chest Radiography, 3) Smear, and 4) Culture. Numbers in overlaps indicate individuals who received multiple diagnostic tests.

Between 2015 and 2018, use of Xpert MTB/RIF and chest radiography diagnostic testing differed significantly between Brazil’s incarcerated and non-incarcerated populations (Fig. 2). Use of Xpert MTB/RIF for incarcerated individuals was greater than that for non-incarcerated individuals (36.2% vs 26.5%, p-value<0.001 each year and overall). Xpert MTB/RIF use in prison settings increased over the four years (p-value<0.001 for a positive linear trend in Xpert MTB/RIF use). Chest radiography use was consistently lower for incarcerated individuals than for non-incarcerated individuals (41.6% vs 74.4%, p-value<0.001 each year and overall).

**Figure 2:**
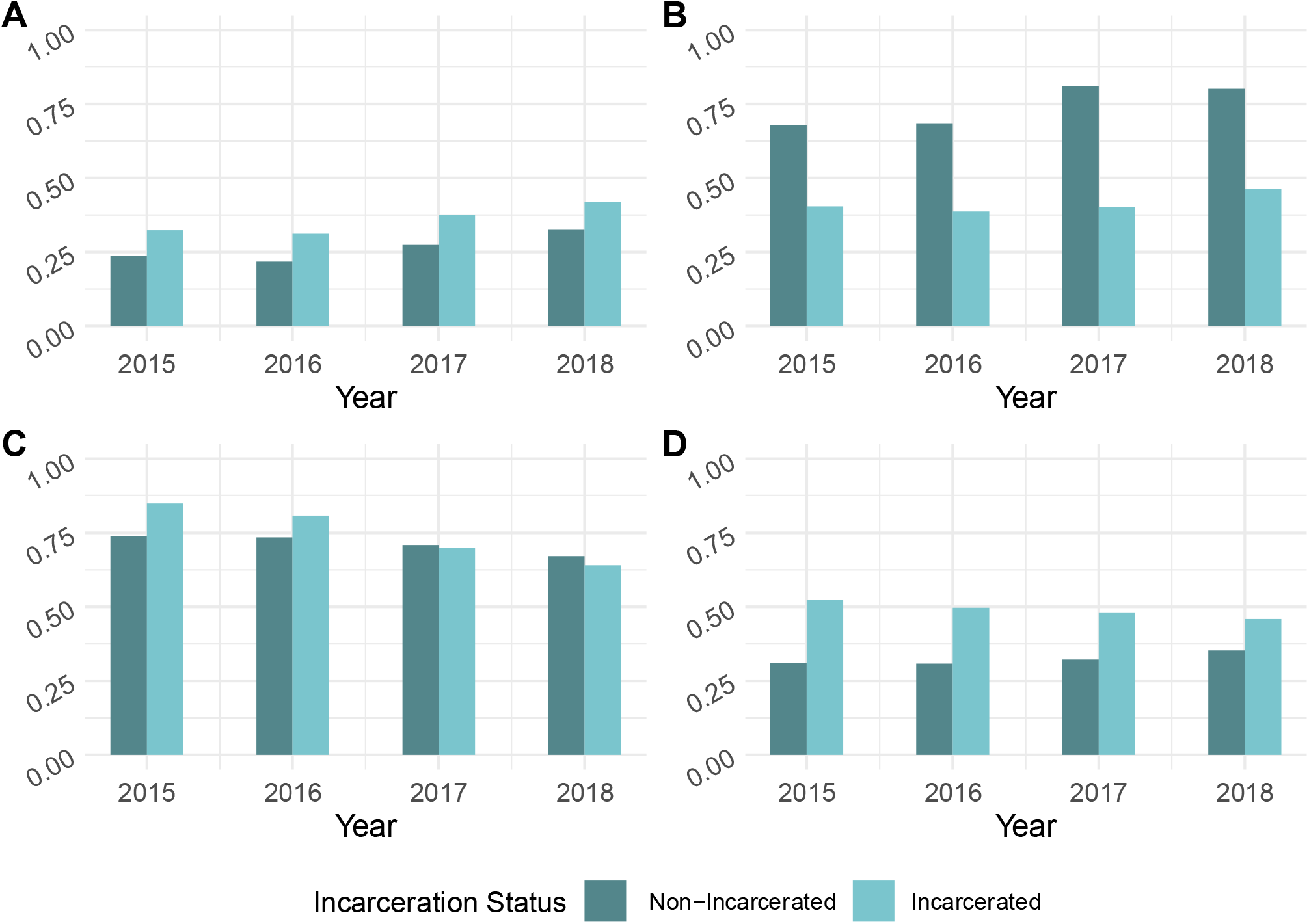
Tuberculosis diagnostic test use in Brazil. Proportions of incarcerated (light blue) and non-incarcerated (dark blue) individuals diagnosed between 2015-2018 by A) Xpert MTB/RIF, B) chest radiography, C) smear, and D) culture.

Across Brazil’s 26 states and one federal district, we found significant heterogeneity in usage in prisons for both Xpert MTB/RIF (X-squared=2717.3, df=26, p<0.001) and chest radiography (X-squared=9039.7, df=26, p<0.001) (Fig. 3). Diagnosis with Xpert MTB/RIF ranged from 4.7% and 4.9% in Pará and Mato Grosso, to 64.5% and 72.4% in Goiás and Amapá, respectively. Similarly, diagnosis with chest radiography ranged from 11.7% and 18.9% in São Paulo and Amapá, to 84.5% and 88.4% in Rio De Janeiro and Espírito Santo, respectively.

**Figure 3:**
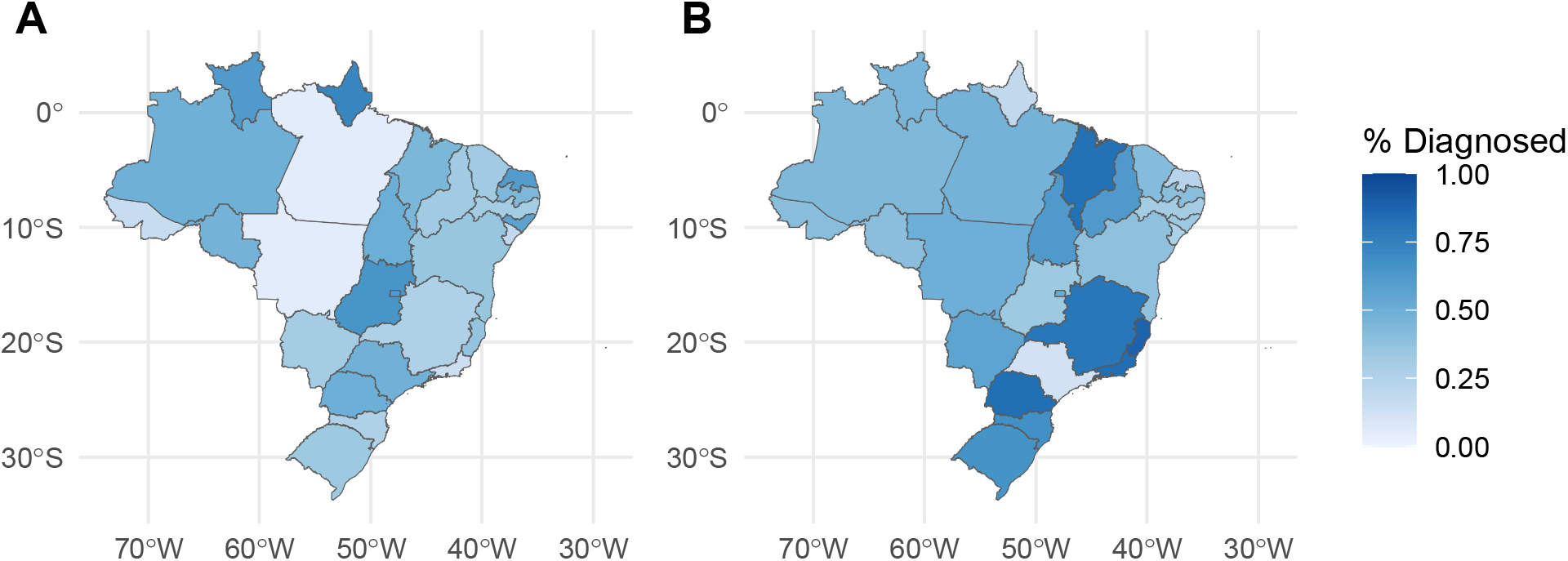
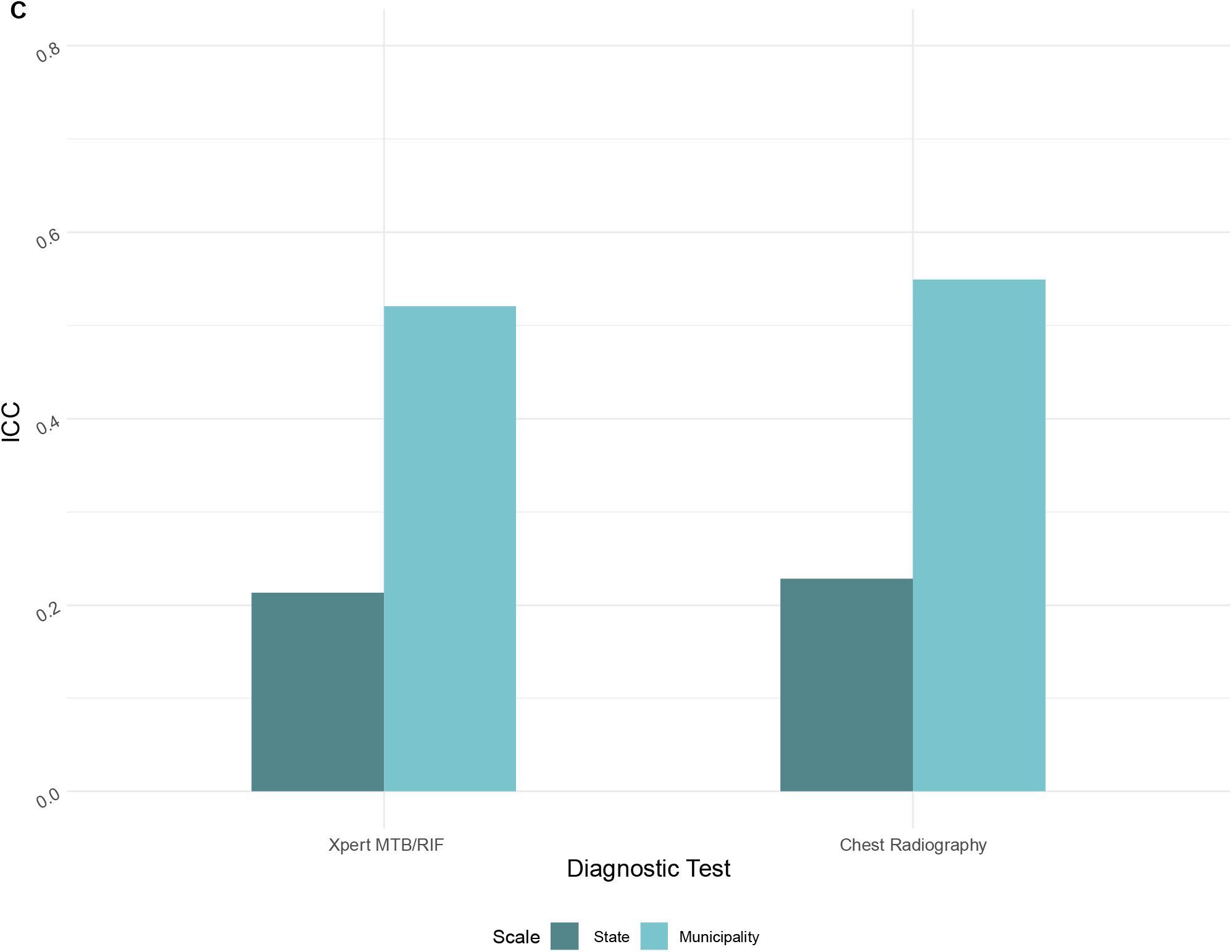
Spatial heterogeneity in the use of Xpert MTB/RIF and chest radiography among the incarcerated population. A) Map of Brazilian states showing the percent of incarcerated individuals diagnosed with Xpert MTB/RIF between 2015 to 2018. Darker colors represent greater diagnostic use. B) Map of Brazilian states showing the percent of incarcerated individuals diagnosed with chest radiography between 2015 to 2018. C) The variance in use of Xpert MTB/RIF and chest radiography attributed to state (dark blue) and municipality (light blue) random effects as measured by the intraclass correlation coefficient (ICC).

Brazil is further divided from the state level into 5,570 municipalities, of which 1,514 contain at least one prison, and 1,235 had at least one TB case from its incarcerated population reported to SINAN between 2015 to 2018. Primary healthcare in Brazil, including healthcare within prisons, is provided by municipalities^25^. We found that 21.3% and 52.1% of variation in Xpert MTB/RIF use, as measured by intraclass correlation coefficients, can be attributed to a patient’s state and municipality, respectively (Fig. 3c). Similarly, 22.8% and 54.9% of variation in chest radiography use among incarcerated populations can be attributed to a patient’s state and municipality, respectively.

To identify other potential predictors of diagnostic use among the incarcerated population, we fit binomial regression models for both diagnostics (Table 1). We found that Xpert MTB/RIF use was greater in 2017 (OR:1.55, CI:1.42-1.69) and 2018 (OR:2.23, CI:2.03-2.44, p<0.001) compared to 2015. A greater municipality incarcerated population size was associated with greater use of Xpert MTB/RIF (OR:1.76, CI:1.49-2.08, p<0.001), though average per capita income, GINI coefficient, and population density were not significantly correlated with Xpert MTB/RIF use. Additionally, we found that GINI coefficient and per capita income were significantly positively associated with chest radiography use, while incarcerated population size was negatively associated with chest radiography use.

**Table 1.** Multivariable regression for use of Xpert MTB/RIF and chest radiography with municipality random effects. The odds ratios (OR), 95% confidence intervals (CI), and p-values are listed for each predictor, with significantly associated covariates in bold.

| *Predictors* | **Access to Xpert MTB/RIF** |  |  | **Access to Chest Radiography** |  |  |
| --- | --- | --- | --- | --- | --- | --- |
|  | *OR* | *95% CI* | *p* | *OR* | *95% CI* | *p* |
| Covariate |  |  |  |  |  |  |
| Population Density* | 1.04 | 0.92 - 1.17 | 0.542 | 1.01 | 0.90 - 1.12 | 0.919 |
| Gini Coefficient** | 0.98 | 0.72 - 1.35 | 0.913 | 1.50 | 1.12 - 2.03 | **0.007** |
| Incarcerated Population Size* | 1.76 | 1.49 - 2.08 | **<0.001** | 0.45 | 0.39 - 0.52 | **<0.001** |
| Average per Capita Income* | 1.40 | 0.88 - 2.22 | 0.156 | 3.33 | 2.10 - 5.30 | **<0.001** |
| Year |  |  |  |  |  |  |
| 2015 | Ref |  |  | Ref |  |  |
| 2016 | 0.97 | 0.88 - 1.06 | 0.458 | 0.89 | 0.80 - 0.99 | **0.026** |
| 2017 | 1.55 | 1.42 - 1.69 | **<0.001** | 0.97 | 0.88 - 1.07 | 0.565 |
| 2018 | 2.23 | 2.03 - 2.44 | **<0.001** | 1.17 | 1.06 - 1.29 | **0.002** |

\*log-scale
\*\*rescaled by a factor of 10

To identify specific gaps in Xpert MTB/RIF and chest radiography usage across Brazil’s incarcerated population, we further examined municipalities with incarcerated populations of at least 5000 individuals and with at least 5 diagnosed TB cases between 2015-2018 (Fig. 4, Table 2). Of these 26 municipalities, seven reported that less than the national average of 36.2% of incarcerated patients were diagnosed with Xpert MTB/RIF, and 18 reported that less than the national average of 41.6% of incarcerated patients diagnosed were with chest radiography.

**Table 2.** Use of TB diagnostics among the incarcerated populations in selected municipalities. We selected municipalities with incarcerated populations of at least 5000 and with at least 5 TB incarcerated TB diagnoses between 2015 to 2018. Table columns indicate the incarcerated population size and TB diagnoses for each municipality, as well as the percent of TB cases diagnosed with Xpert MTB/RIF or chest radiography, from 2015-2018.

| Region | State | Municipality | Incarcerated Population Size, 2018 | Incarcerated TB Diagnoses, 2015-2018 | % Diagnosed with Xpert MTB/RIF | % Diagnosed with Chest Radiography |
| --- | --- | --- | --- | --- | --- | --- |
| North | RO | Porto Velho | 6475 | 265 | 55.1 | 37.7 |
|  | AM | Manaus | 7748 | 311 | 53.4 | 39.5 |
|  | PA | Santa Izabel Do Pará | 5439 | 339 | 1.5 | 40.4 |
| Northeast | MA | São Luís | 5462 | 305 | 59.3 | 86.9 |
|  | CE | Itaitinga | 9670 | 569 | 38.0 | 24.4 |
|  | PB | João Pessoa | 5203 | 214 | 53.1 | 33.9 |
|  | PE | Recife | 6409 | 560 | 34.1 | 20.0 |
|  | AL | Maceió | 7619 | 107 | 67.3 | 25.2 |
|  | BA | Salvador | 5001 | 209 | 67.9 | 31.1 |
| Southeast | MG | Ribeirão Das Neves | 9522 | 34 | 0.0 | 82.4 |
|  |  | São Joaquim De Bicas | 5278 | 17 | 5.9 | 47.1 |
|  | ES | Vila Velha | 7958 | 18 | 16.7 | 72.2 |
|  | RJ | Rio De Janeiro | 36243 | 3417 | 13.8 | 83.8 |
|  | SP | Bauru | 6601 | 347 | 65.1 | 16.4 |
|  |  | Franco Da Rocha | 9804 | 317 | 12.3 | 14.8 |
|  |  | Guarulhos | 7457 | 380 | 54.5 | 3.9 |
|  |  | Hortolândia | 6741 | 307 | 64.2 | 5.2 |
|  |  | Lavínia | 6291 | 297 | 63.0 | 2.0 |
|  |  | Mirandópolis | 5404 | 201 | 60.7 | 13.9 |
|  |  | São Paulo | 15252 | 914 | 65.0 | 21.0 |
|  |  | Tremembé | 6837 | 187 | 62.6 | 2.1 |
| South | RS | Charqueadas | 5142 | 240 | 67.1 | 25.0 |
|  |  | Porto Alegre | 6204 | 614 | 57.8 | 92.0 |
| Central- | MS | Campo Grande | 5572 | 435 | 37.9 | 49.0 |
| West | GO | Aparecida De<br>Goiânia | 6440 | 268 | 87.3 | 6.0 |
|  | DF | Brasília | 16359 | 138 | 60.9 | 51.4 |

**Figure 4:**
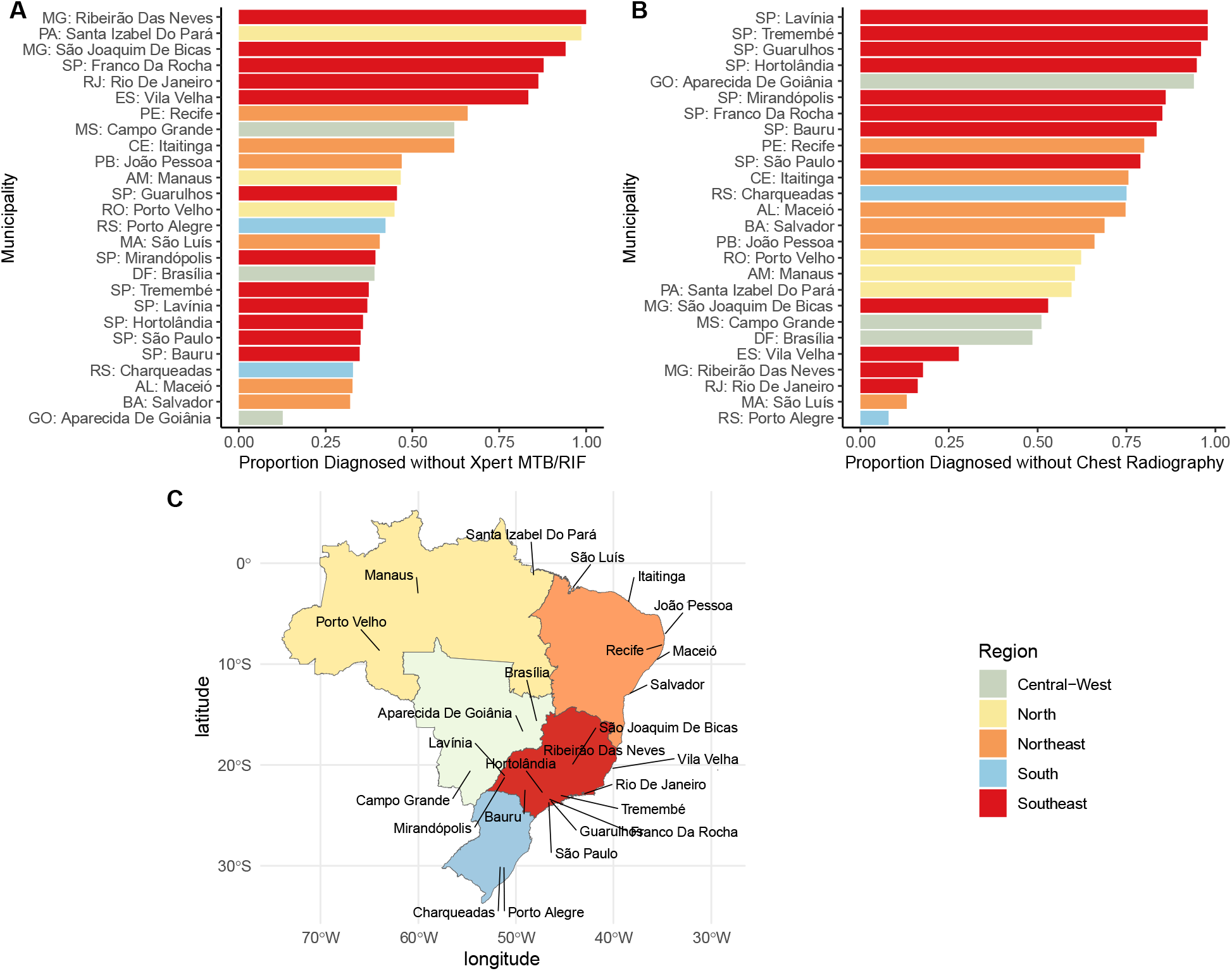
Gaps use of Xpert MTB/RIF and chest radiography in the incarcerated population. A) The proportion of incarcerated individuals in each municipality who were diagnosed without Xpert MTB/RIF, colored by region. Longer bars suggest a greater gap in diagnostic use. B) The proportion of incarcerated individuals in each municipality who were diagnosed without chest radiography, colored by region. C) The 26 municipalities labeled within a map of Brazil, colored by region.

## DISCUSSION

We found that the use of TB diagnostic tests differs significantly between the incarcerated and general populations in Brazil. The use of Xpert MTB/RIF was greater among the incarcerated population and expanded in 2017 and 2018 compared to 2015, as expected with the gradual implementation of this test across prisons. Further, we found that diagnosis with Xpert MTB/RIF and chest radiography is largely determined by a patient’s state and municipality at time of diagnosis, reflecting the varying TB programs and control efforts organized by Brazil’s municipalities. Finally, we identified several gaps in which municipalities with large prison populations had very limited use of Xpert MTB/RIF and/or chest radiography.

As a significant and increasing proportion of TB notifications in Brazil occur among the incarcerated population^9^, and as prisons can amplify community TB epidemics^13–15^, targeting prisons for TB control efforts can substantially impact Brazil’s overall TB incidence. Given the high accuracy of Xpert MTB/RIF in diagnosing TB and its effectiveness within prison settings^6,16,26,27^, ensuring equitable access to this diagnostic across Brazil’s incarcerated population is critical. Our findings suggest that efforts to expand Xpert MTB/RIF availability in prisons have been effective, as a greater proportion of incarcerated versus non-incarcerated individuals were diagnosed with this test, and as the proportion diagnosed increased through the study period. Yet our observation of significant spatial heterogeneity in Xpert MTB/RIF use across Brazil’s prisons indicates that a large population in Brazil may not currently have access to this tool based on where they live. Thus, targeting the specific municipalities identified as having large incarcerated populations and limited Xpert MTB/RIF use should be a focus of continued Xpert MTB/RIF expansion and overall TB control efforts as addressing issues in these settings could help to further limit TB incidence in and beyond prison settings.

Reducing the TB diagnostic gap remains a global challenge, and efforts to expand access to Xpert MTB/RIF testing in prisons and across healthcare systems globally will be critical to achieve the WHO End TB Strategy of an 80% decrease in new cases by 2030^4^. Some countries have reported rapid expansion of Xpert MTB/RIF. For example, in Uzbekistan, Xpert MTB/RIF coverage increased from 24% in 2018 to 46% in 2019^28^, and in El Salvador, the use of Xpert MTB/RIF testing as part of a package of TB diagnostic interventions tripled incarcerated TB notifications in five years^29^. However, barriers to equitable diagnostic access remain, such as diagnostic machine service coordination issues^30^, and further research will be necessary to identify diagnostic gaps and policy solutions. Our findings of significant spatial heterogeneity in Xpert MTB/RIF usage across Brazil and by incarceration status highlight the importance of understanding diagnostic availability for other national TB control programs.

There are several limitations to this study. First, availability of specific diagnostic tests across Brazil’s diagnostic facilities is not centrally reported, and individual access to diagnostic tests cannot be ascertained from SINAN. We investigated the usage of different diagnostics among confirmed cases, which may reflect availability of diagnostics in a particular setting, functionality, and selection by healthcare providers. However, we were unable to investigate how widely Xpert MTB/RIF is used among all individuals with suspected TB, and hence cannot report usage for individuals who did not actually receive a confirmed diagnosis. Another limitation was the use of 2010 socioeconomic indicators in the binomial regression. Though these values have been improving over time, the 2015 economic crisis could have variably influenced socioeconomic factors across municipalities; as a census has not been conducted since, the 2010 data still provide a strong estimate. Third, we excluded cases where the incarceration status was not reported. This represents under 3.2% of total cases (9,086 of 284,415), so we have confidence that excluding diagnostic usage data for this population does not significantly impact our results. Finally, our study focuses on years preceding the COVID-19 pandemic. Understanding the effect of COVID-19 on widening TB diagnostic gaps within and outside prisons is critical for TB control programs^1^.

Ultimately, to achieve the WHO End TB goals, targeted expansion of rapid molecular diagnostics for incarcerated populations and regions with limited diagnostic usage will be essential and could reduce significant diagnostic gaps.

## Data Availability

All data produced in the present study are available upon reasonable request to the authors

## Funding

National Institutes of Health grant R01 AI130058 (JRA) and R01 AI149620 (JRA and JC).

EBF, KSW, JRA, and JC conceived of the study. EBF analyzed data and made data visualizations. KSW and JC acquired data. All authors contributed to paper revisions and approved of the final version.

## Notes

### Competing Interest Statement

The authors have declared no competing interest.

### Author Declarations

This study was approved by the Institutional Review Board at Stanford University (Protocol #50466).

